# Identifying Mitral Valve Prolapse at Risk for Ventricular Arrhythmias and Myocardial Fibrosis from 12-lead Electrocardiograms using Deep Learning

**DOI:** 10.1101/2021.12.23.21268341

**Authors:** Geoffrey H. Tison, Sean Abreau, Lisa Lim, Valentina Crudo, Joshua Barrios, Thuy Nguyen, Gene Hu, Shalini Dixit, Gregory Nah, Yoojin Lee, Francesca N. Delling

## Abstract

**Background:** Mitral valve prolapse (MVP) is a common valvulopathy, with a subset of MVP patients developing sudden cardiac death or cardiac arrest. Complex ventricular ectopy (ComVE) represents a marker of arrhythmic risk that is associated with myocardial fibrosis and increased mortality in MVP. We hypothesize that an ECG-based machine-learning model can identify MVP with ComVE and/or myocardial fibrosis on cardiac magnetic resonance (CMR) imaging.

**Methods:** A deep convolutional neural network (CNN) was trained to detect ComVE using 6,916 12-lead ECGs from 569 MVP patients evaluated at the University of California San Francisco (UCSF) between 2012 and 2020. A separate CNN was also trained to detect late gadolinium enhancement (LGE) using 87 ECGs from MVP patients with contrast CMR. **Results**: The prevalence of ComVE was 160/569 or 28% (20 patients or 3% had cardiac arrest or sudden cardiac death). The area under the curve (AUC) of the CNN to detect ComVE was 0.81 (95% CI, 0.78-0.84). AUC remained high even after excluding patients with moderate-severe mitral regurgitation (MR) [0.80 (95% CI, 0.77-0.83)], or with bileaflet MVP [0.81 (95% CI, 0.76-0.85)]. The top ECG segments able to discriminate ComVE vs no ComVE were related to ventricular depolarization and repolarization (early-mid ST and QRS fromV1, V3, and III). LGE in the papillary muscles or basal inferolateral wall was present in 21 (24%) of 87 patients with available CMR. The AUC for detection of LGE was 0.75 (95% CI, 0.68-0.82).

**Conclusions:** Standard 12-lead ECGs analyzed with machine learning can detect MVP at risk for ventricular arrhythmias and fibrosis and can identify novel ECG correlates of arrhythmic risk regardless of leaflet involvement or mitral regurgitation severity. ECG-based CNNs may help select those MVP patients requiring closer follow-up and/or a CMR.

## INTRODUCTION

Mitral valve prolapse (MVP) is a common valvulopathy affecting over 170 million worldwide.^1, 2^ Although a subset of MVP patients (0.14-1.8% yearly)^3-9^ will develop cardiac arrest (SCA) or sudden cardiac death (SCD), and 7% of SCDs in the young are caused by MVP,^10^ predictors of this devastating outcome are not well defined. To date, it has been challenging to pinpoint a unique ECG phenotype of SCD/SCA in MVP. Inverted or biphasic T waves have been described in the inferior ECG leads with a prevalence ranging between 30 and 78% in selected, retrospective studies of patients with SCD/SCA.^5, 10^ However, inferior T wave abnormalities are present in 40% of MVPs even if no history of ventricular arrhythmia.^11^ QT prolongation and QT dispersion in malignant MVP syndrome have been described in some, but not all, studies.^12-14^ An easily obtainable and fully automated “surveillance” tool for SCD/SCA in MVP would provide substantial clinical value to quickly identify those at higher arrhythmic risk within large clinical cohorts of mostly benign MVPs.

Previously, SCD/SCA in MVP has been linked to flail leaflet and severe mitral regurgitation (MR).^9^ Other studies have reported a high arrhythmic risk in a bileaflet phenotype with mild MR, T wave abnormalities in the inferior ECG leads, and complex ventricular ectopy [ComVE - defined as frequent polymorphic premature ventricular contractions (PVCs), bigeminy, or non-sustained ventricular tachycardia (NSVT)] (**Figure 1**).^5, 10, 15^ In the bileaflet phenotype, often associated with annular disjunction leading to abnormal annular mechanics and localized traction on the myocardium, autopsies and cardiac magnetic resonance (CMR) imaging with late gadolinium enhancement (LGE) have revealed focal (replacement) fibrosis in the papillary muscles or inferolateral base of the left ventricle.^16, 17 10, 18, 19^ However, in imaging studies of living individuals and in post-mortem samples of malignant MVP cases, bileaflet involvement and focal fibrosis are not consistent findings.^10 20, 21 22, 23^ Regardless of leaflet involvement or degree of MR, ComVE is detected in 80-100% of MVPs prior to SCA or SCD. ComVE is associated with myocardial fibrosis and is linked to higher all-cause mortality.^5, 10, 24^ Ambulatory ECG monitoring to detect ComVE may not be available for all MVP patients, especially if asymptomatic.

**Figure 1.**
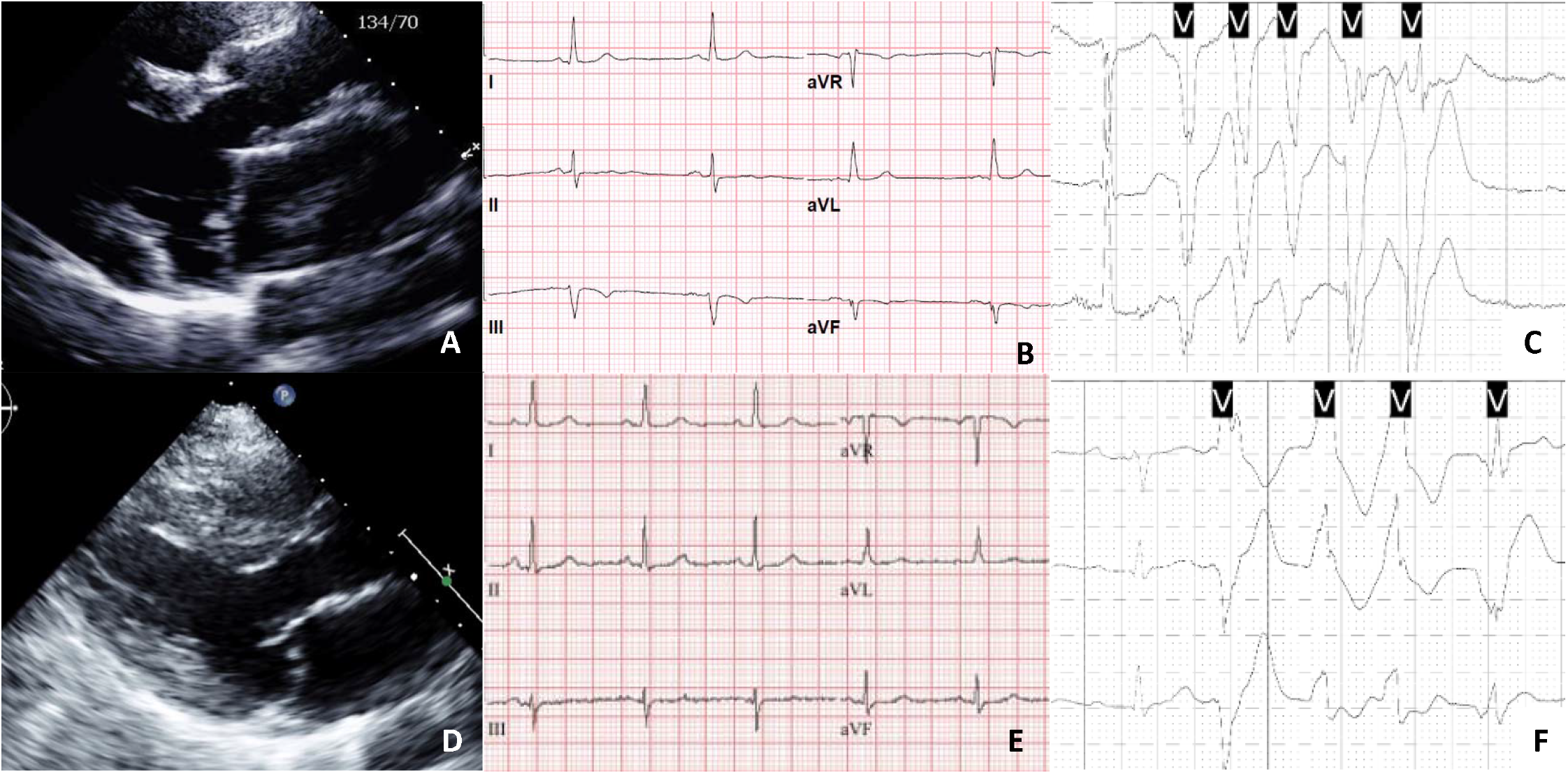
Echocardiographic example of mitral valve prolapse (MVP) with bileaflet involvement (A) and inverted or biphasic T wave inversions in the inferior ECG leads (B). Posterior MVP (D) without repolarization abnormalities on ECG (E). Both MVP cases had complex ventricular ectopy (C and F) prior to experiencing a cardiac arrest.

We have previously demonstrated that machine learning-based methods can be applied to accurately analyze ECGs to discriminate patients with and without MVP.^25^ Here we investigate whether ECGs can discriminate the “arrhythmic” from the “non-arrhythmic” MVP phenotype. Specifically, we hypothesize that an ECG-based deep-learning model can 1) identify MVP at risk for ComVE, including those that will develop sustained VT or ventricular fibrillation 2) identify novel ECG correlates of MVP-related myopathy and arrhythmic risk beyond traditional ECG criteria, and across mono or bileaflet MVP subtypes and 3) select those MVPs at risk for myocardial fibrosis on CMR.

## METHODS

### Study population

We identified 638 consecutive MVP cases between January 2012 and December 2020 who had at least one ECG and echocardiogram at UCSF. Nine patients were excluded because of concomitant arrhythmogenic conditions: hypertrophic cardiomyopathy (2), sarcoidosis (2), Wolff-Parkinson-White (1), ischemic (2) and non-ischemic (2) cardiomyopathy with LV ejection fraction ≤ 35% (**Figure 2**). An additional 60 patients were excluded because of lack of ECGs or presence of atrial fibrillation, PVCs, paced rhythms or right/left bundle branch on ECG (**Figure 2**).

**Figure 2.**
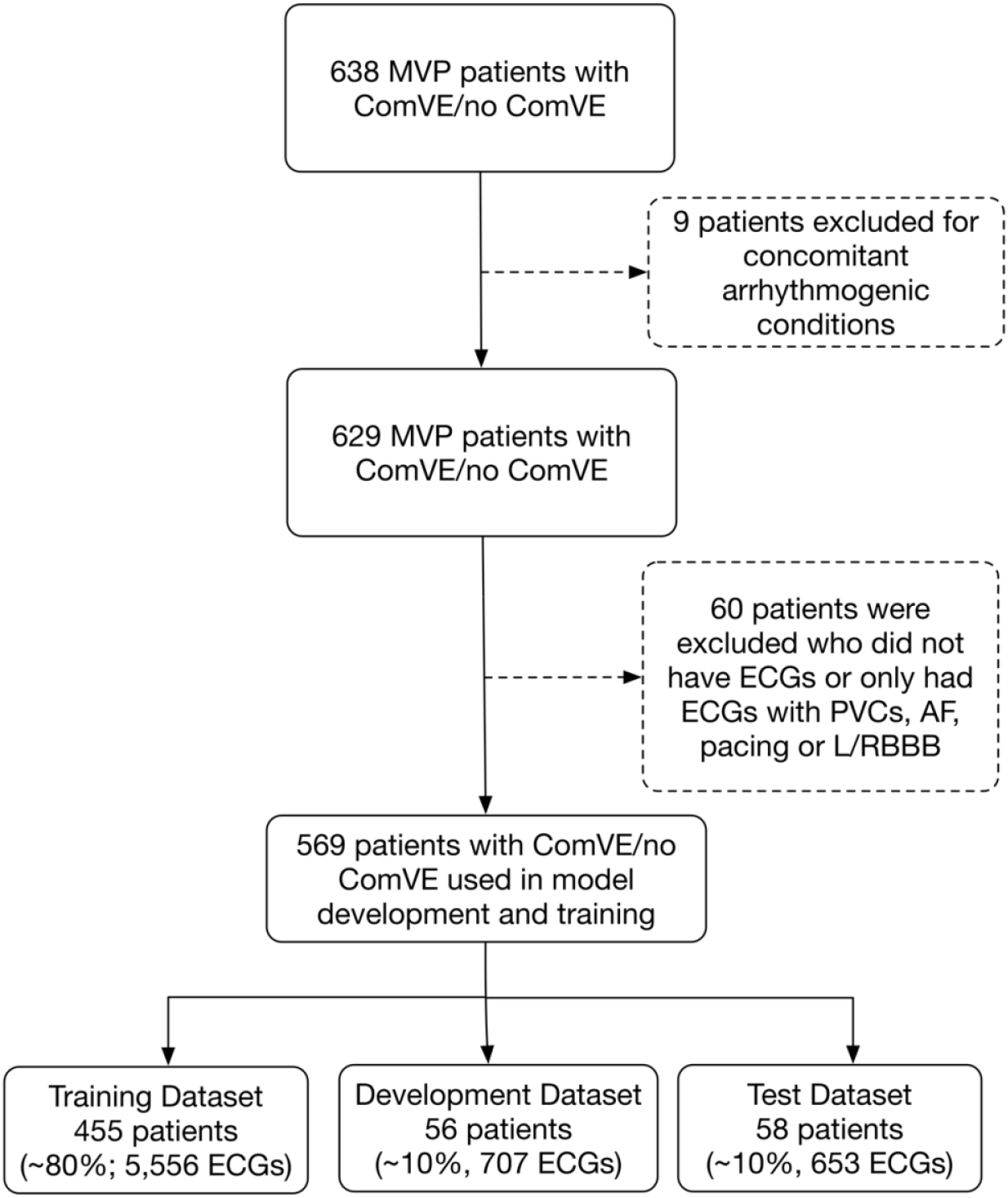
Diagram of study cohorts and datasets. UCSF patients with a confirmed diagnosis of complex ventricular ectopy (ComVE) or no ComVE were selected for analysis. Patients meeting criteria were randomly split into training, development, and test datasets in a ratio of approximately 8:1:1. The development dataset was used to monitor performance during convolutional neural network (CNN) training. The test dataset was used to evaluate model performance on unseen data. MVP = mitral valve prolapse; ECG = electrocardiogram; PVCs = premature ventricular contractions; AF = atrial fibrillation; L/RBBB = left/right bundle branch block

All remaining 569 MVP subjects had demographic, clinical, ECG, echocardiographic, and (if available) CMR study data already extracted and organized in an online research database as part of a UCSF MVP Registry. UCSF Institutional Review Board approval was obtained for this study. Patient’s informed consent was exempted by the Institutional Review Board because of the retrospective nature of the study.

ECG data was extracted in XML format for 12-leads over 10 seconds, recorded at either 250Hz or 500Hz. All 500Hz ECG records were down-sampled to 250Hz, and all ECG signal data was represented as a 2500×12 matrix.

### Cardiac Imaging

MVP was defined as systolic leaflet displacement of one or both leaflets >2 mm beyond the mitral annulus in a parasternal or apical 3-chamber long-axis echocardiographic view (**Figure 1**). When quantitative assessment of MR was not available, its severity was based on visual estimation of the regurgitant jet. LV end-diastolic/end-systolic volumes, ejection fraction, and mass were quantified and indexed to body surface area. Right ventricular dilatation was defined as a basal diameter ≥ 4.2 cm in the 4-chamber view. Right ventricular systolic dysfunction was assessed qualitatively.

CMR was conducted through the UCSF CMR Core using a 3.0T Discovery MR750w scanner (GE Healthcare, Milwaukee, WI). Breath-hold cine imaging was performed using a segmented balanced steady state free precession pulse sequence to assess left/right ventricular structure and function in short and long-axis views. Subjects received a bolus IV injection of 0.1 mmol/kg gadobutrol (Gadavist). Short and long axis plane high-resolution phase sensitive inversion recovery delayed enhancement images were obtained 10 min minutes after contrast injection for detection and quantification of myocardial scar.

### Identification of study groups

We extracted raw ECG voltage data for study patients in the UCSF MVP registry who were classified into 2 groups: patients with ComVE and without ComVE. ComVE was defined as: greater than 5% burden of PVCs (isolated or in bigeminy) or presence of NSVT on 48-hour Holter or 2-week event monitor.^5^ NSVT was defined as ≥ 3 PVCs with a rate >100 bpm lasting <30 seconds.^15^ In addition to those with ambulatory ECG monitoring, the ComVE or arrhythmic group also included MVP cases with 1) ComVE on inpatient telemetry strips or 2) a secondary prevention implantable cardioverter defibrillator (ICD) following a cardiac arrest due to ventricular fibrillation or hemodynamically significant sustained VT (spontaneous or induced during EP study) or 3) sustained VT (spontaneous or induced) treated with radiofrequency catheter ablation. Sustained VT was defined as tachycardia of ventricular origin with a rate >100 bpm and lasting >30 seconds.^15^

The non-ComVE group included MVP patients that 1) had ambulatory ECG or telemetry monitoring but no evidence of ComVE (< 5% or no PVCs and absence of NSVT) and/or 2) did not have an international classification of disease (ICD) 9 or 10 codes for VT, PVCs, cardiac arrest, or a current procedural terminology (CTP) code for ICD placement and/or 3) did not have a complaint of palpitations on review of medical records.

Given the known association of MVP with myocardial fibrosis,^10, 18, 23^ we performed a separate analysis classifying all MVP patients regardless of ComVE status into 2 groups: patients with and without LGE on contrast CMR.

In both analyses, only ECGs following the first echocardiographic diagnosis of MVP were included. The data was divided into training (80%), development (10%) and testing (10%) datasets, split randomly by patient. All ECGs from patients with or without ComVE or LGE were given a respective label and used for algorithm training and testing. In sensitivity analysis, restricting analysis to ECGs within one year of the date of diagnosis did not materially affect results.

### Algorithm Development

We trained separate CNNs to predict ComVE and LGE from a 12-lead ECG voltage matrix of size 2,500 x12. The final output of the CNN for each task correlates with disease probability and is referred to as the ComVE or LGE score. The model had a one-dimensional ResNet architecture.^26, 27^ Our CNN was initialized from a pretrained model trained on 38 arrhythmias and 992,748 ECGs obtained from UCSF between 2003-2018 as previously described,^28^ keeping all layers trainable. The model output consists of two mutually exclusive binary classes, each with a probability value between 0 and 1. It contained 34 stacked convolutional layers with filter size of eight, each with a convolution, a ReLU non-linearity,^29^ and a batch normalization layer.^30^ Every other layer is connected by a residual layer and every 4 convolutional layers a max-pooling layer is applied, which reduces the time resolution by a factor of 2 starting from 2500. The number of convolutional channels is doubled every 8 layers starting from 64. The first and last layer differ from the others due to placement of activations and the output of the final layer of convolution is a 256 × 10 matrix which then feeds into a sigmoid activation function. This output between 0-1 for each ECG is referred to as the CNN score which is correlated with the predicted probability of the outcome. The Adam optimizer trained our model until convergence with a learning rate of 10^−4^, then the learning rate was reduced by a factor of 10 and training continued to convergence twice.^31^ We searched over initial learning rates from 10^−6^ to 10^−3^. We used TensorFlow version 1.14 and Python 2.7.

### Examining ECG-segments most strongly contributing to predictions

To examine the ECG segments contributing most strongly to the ability to predict ComVE or LGE, we trained models using a parallel machine learning approach designed for increased interpretability, as previously reported.^25^ In brief, we applied a trained CNN model to segment ECGs into standard segments and intervals (P wave, PR, QRS, ST), sampled a 725-length vector representation for every ECG based on its segmentation, and trained a gradient boosted model for classification.^25^ Identical cohorts and training datasets as the primary analysis were used for this approach. Variable importance scores were then derived from separate models trained for ComVE and LGE, demonstrating the ECG segments contributing most strongly to each model prediction. To increase the robustness of ECG-segment variable importance estimates, 10 models were separately trained for each task, using random seeds, and we reported variable importance scores averaged over the 10 models.

### Statistical Analysis

We compared MVP subjects with ComVE vs non-ComVE with regard to demographics, comorbidities, ECG, echocardiographic variables, and, when CMR studies were available, presence of LGE. Continuous variables were expressed as mean and standard deviations. Categorical data were expressed as number and percentage of total subjects in each group. Differences between the 2 groups were assessed using the chi-square test for categorical variables and t-test for continuous variables. Analyses used Stata Version 14.2 (StataCorp LP). A 2-tailed *P* < 0.05 was considered statistically significant.

In the holdout test dataset for each analysis, CNN performance was evaluated by calculating the area under the receiver operating characteristic curve (AUC), sensitivity, specificity, positive predictive value (PPV), negative predictive value (NPV) and F1 score (harmonic mean of the PPV and sensitivity) compared to the gold-standard definitions of ComVE or LGE as described above. A threshold for binary decision was chosen to optimize the F1 score to predict ComVE (threshold 0.39) in the validation dataset and then applied to the test set for calculation of the F1 score, sensitivity and specificity and predictive values. Confidence intervals for AUCs were computed using the bootstrap method using 95% of the data over 200 samples. Binary thresholds to evaluate performance for each class were chosen to maximize the geometric mean of the positive predictive value and sensitivity in the development dataset for that analysis.

## RESULTS

In the cohort of 569 patients with MVP (160 or 28% with ComVE), we analyzed a total of 6916 ECGs. **Table 1** summarizes the baseline characteristics for the ComVE and non-ComVE groups. The 2 groups had similar demographics, prevalence of cardiovascular risk factors and coronary artery disease. Atrial fibrillation and flutter were more prevalent in the ComVE group.

**Table 1.**
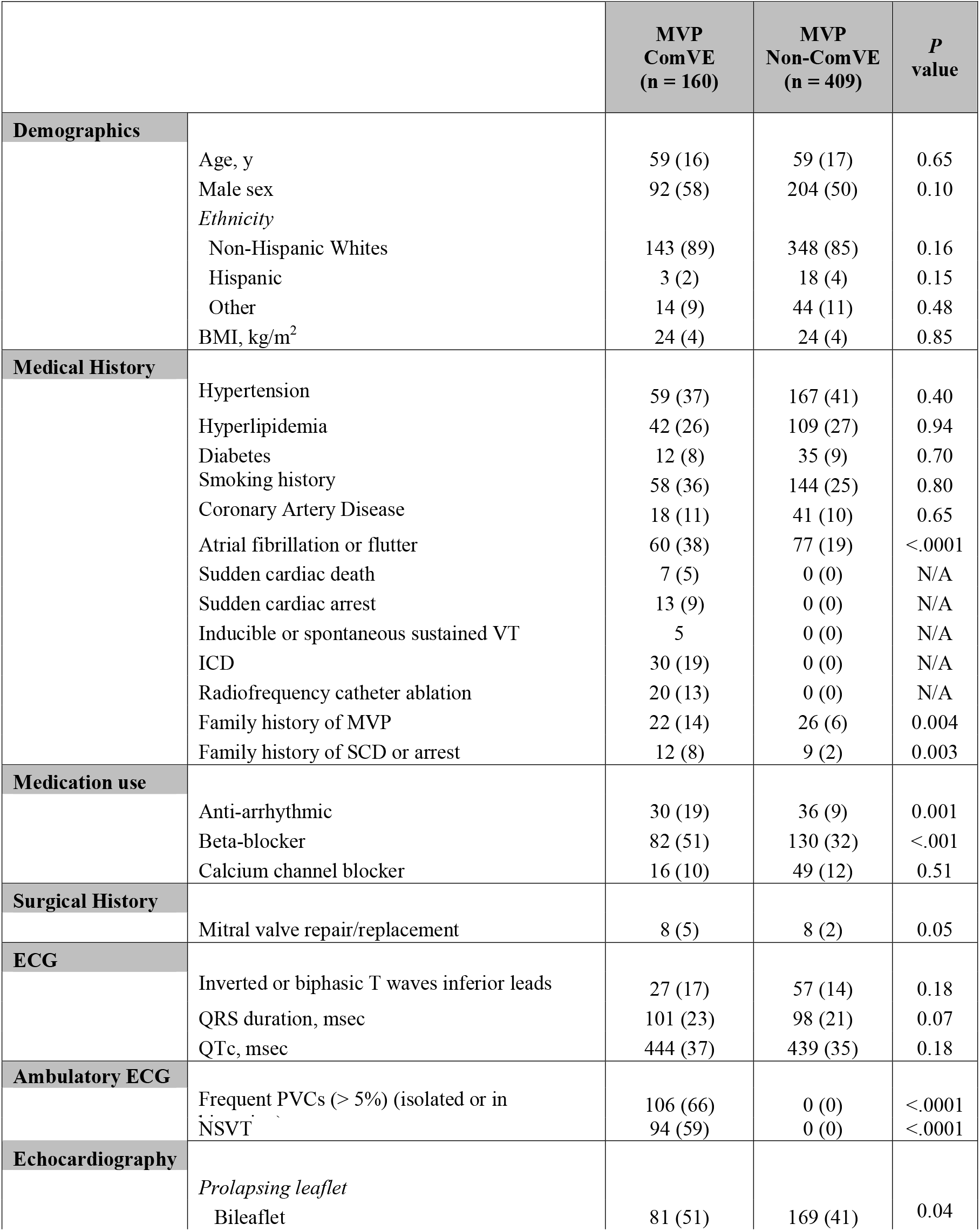
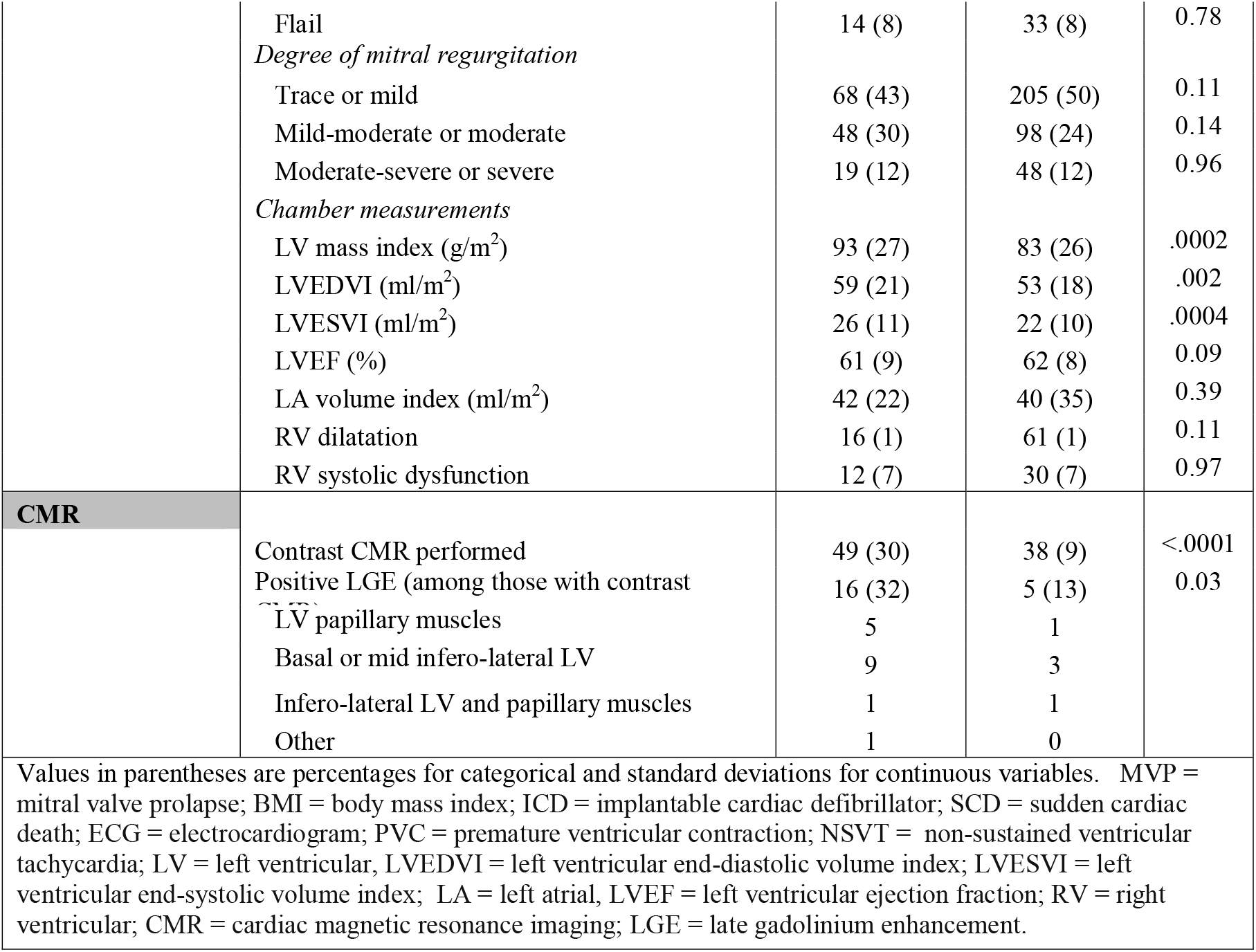
Baseline characteristics of MVP patients in the testing cohort according to the presence or absence of complex ventricular ectopy (ComVE).

Despite greater use of antiarrhythmics and beta-blockers among those with ComVE (**Table 1**), 20 individuals (12% of the ComVE group, 3% of the total MVP sample) experienced SCD or an aborted cardiac arrest. Baseline ECG characteristics, including the presence of inverted or biphasic T waves, did not differ between those with and without ComVE. There was a higher proportion of patients with bileaflet involvement and LGE among those with ComVE. Compared to the non-ComVE group, LV volumes were larger despite similar LV systolic function and degree of MR.

### Performance of a CNN to predict ComVE

In the test dataset for ComVE, the CNN had an area under the receiver operating characteristic curve (AUC) to detect ComVE of 0.81 (95% CI, 0.78-0.84) (**Figure 3A**) and F1 score of 0.77 (**Table 2**); sensitivity was 78.3%, specificity was 71.3%, PPV 75.9%, NPV 74.0% (**Figure 3B, Table 2**). In patients with ComVE compared to those without ComVE, the ComVE CNN score was higher overall (**Figure 4A**). Within the ComVE group, patients who progressed to severe ventricular arrhythmia (i.e. those with high PVC burden requiring radiofrequency catheter ablation or with sustained VT or ventricular fibrillation arrest/ICD or with SCD) had a higher overall ComVE CNN score compared to ComVE patients who did not progress (**Figure 4B**). When plotted over time from the date of ComVE diagnosis, ComVE CNN scores of MVPs without ComVE were mostly below the ComVE threshold, while scores of ComVE MVPs were generally above (**Figure 4C**). Interestingly, those with ComVE with progression exhibited an increase in CNN score over time, while those without progression exhibited an overall flat to slight downtrend in CNN score. MVPs with ComVE (either with or without progression) had an early drop in ComVE CNN score within the first third of total follow-up time after ComVE diagnosis (**Figure 4C**). Among those with ComVE, initiation of nodal agents or antiarrhythmic medications was more common in the first third of total follow-up time after ComVE diagnosis compared to before ComVE diagnosis (19% versus 6%, respectively, *P*<0.05), which may explain the decrease in ComVE CNN score. MVP patients without ComVE did not exhibit the same early drop in ComVE CNN score, and the use of nodal agents or antiarrhythmics was less common in this group (**Table 1**).

**Figure 3.**
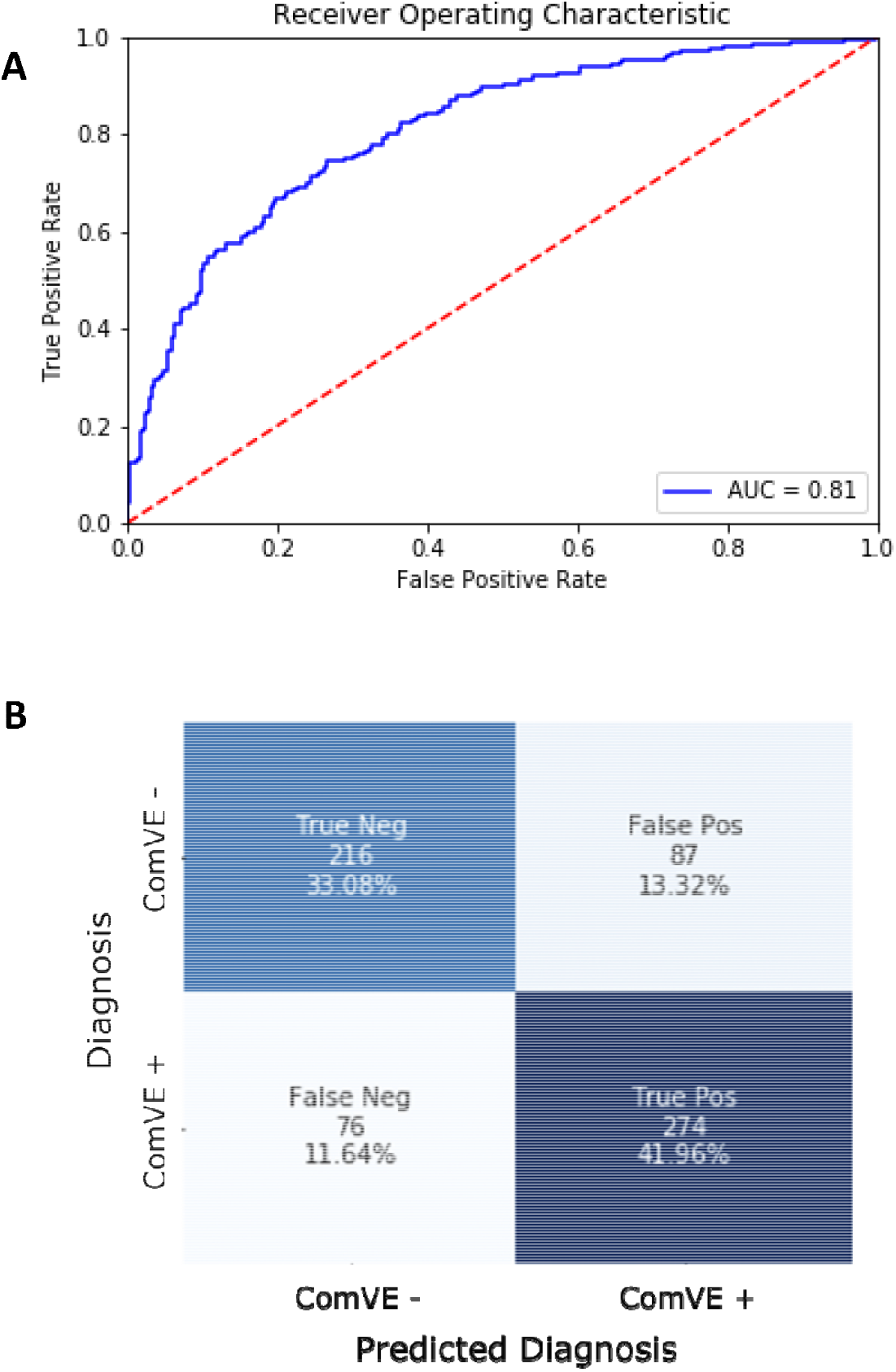
Performance of the convolutional neural network to detect complex ventricular ectopy (ComVE) in the test dataset. (A) Receiver operating characteristic curve for the convolutional neural network (CNN) to predict ComVE. AUC = Area Under the receiver operating characteristic curve. (B) Confusion matrix demonstrating CNN performance to predict ComVE by ECG in the hold-out test dataset at the chosen score threshold of 0.39.

**Table 2.** Performance of the convolutional neural network on the main cohort and on sensitivity analysis cohorts with specified exclusions.

| COHORT | AUC | SENSITIVITY | SPECIFICITY | POSITIVE<br>PREDICTIVE<br>VALUE | NEGATIVE<br>PREDICTIVE<br>VALUE | FALSE<br>POSITIVE<br>RATE | ACCURACY | F1 SCORE |
| --- | --- | --- | --- | --- | --- | --- | --- | --- |
| **MAIN ANALYSIS** |  |  |  |  |  |  |  |  |
| ComVE | 0.81<br>(0.78-0.84) | 0.78<br>(0.70 – 0.83) | 0.71<br>(0.62-0.80) | 0.76<br>(0.70-0.82) | 0.74<br>(0.70-0.78) | 0.29<br>(0.18-0.41) | 0.75<br>(0.71-0.80) | 0.77<br>(0.73-0.81) |
| **SENSITIVITY**<br>**ANALYSES** |  |  |  |  |  |  |  |  |
| Excluded<br>MV Repair or<br>Replacement | 0.81<br>(0.79-0.84) | 0.64<br>(0.57 - 0.76) | 0.87<br>(0.79-0.95) | 0.67<br>(0.57-0.78) | 0.85<br>(0.78-0.92) | 0.13<br>(0.03-0.24) | 0.80<br>(0.74-0.85) | 0.65<br>(0.58-0.72) |
| Excluded<br>Moderate-Severe<br>MR | 0.80<br>(0.77-0.83) | 0.83 (0.71-0.95) | 0.55<br>(0.43-0.67) | 0.59<br>(0.44-0.74) | 0.81<br>(0.74-0.88) | 0.45<br>(0.29-0.53) | 0.67<br>(0.61-0.73) | 0.70<br>(0.65-0.75) |
| Excluded<br>Bileaflet MVP | 0.81<br>(0.76-0.85) | 0.87 (0.81-0.93) | 0.60<br>(0.51-0.69) | 0.76<br>(0.67-0.85) | 0.75<br>(0.67-0.83) | 0.40<br>(0.34-0.52) | 0.76 (0.68-0.81) | 0.81<br>(0.73-0.89) |

AUC = area under the receiver operating characteristic curve, ComVE = Complex ventricular ectopy, MV = mitral valve, MR = mitral regurgitation; MVP = mitral valve prolapse. In parentheses, 95% confidence intervals.

**Figure 4.**
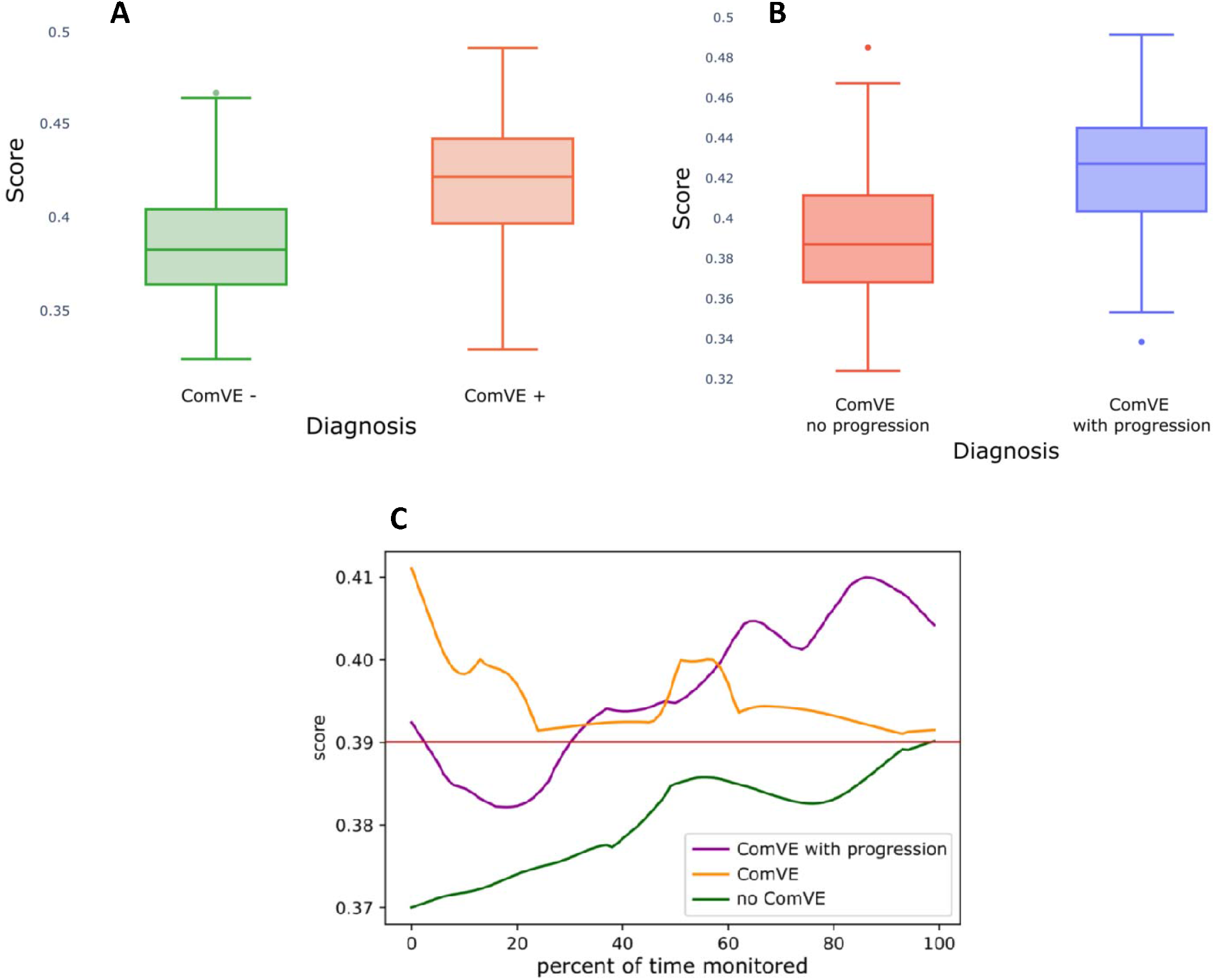
Distribution of ComVE CNN scores by strata in the test dataset. (A) Box and whisker plots showing distributions of ComVE CNN scores for mitral valve prolapse (MVP) patients with and without ComVE. (B) Box and whisker plot showing distributions of ComVE CNN scores for ComVE patients with and without progression. (C) Averaged ComVE CNN scores by patient strata plotted over time, as a percent of total follow-up time since ComVE diagnosis (x-axis). The maximal follow-up time in this cohort was 3220 days (8.8 years). For ComVE patients only (with or without progression), averaged ComVE CNN scores exhibited a drop within the first third of follow-up time. The red line indicates CNN score threshold used for binary classification of ComVE.

We conducted multiple sensitivity analyses as a means to better understand the pathophysiologic correlates of the CNN’s ability to predict ComVE from ECG alone. We separately excluded patients from the dataset who had a history of MV repair or replacement, moderate-severe or greater MR, or bileaflet MVP, and examined the ability of the trained CNN to discriminate ComVE. The degree to which the trained CNN performance remained robust to these exclusions suggested that the CNN identified ComVE independently from the influence of these conditions (**Table 2**). For each sensitivity analysis performed, the AUCs remained similar to that of the primary analysis. Since the same threshold was applied in each sensitivity analysis as the primary analysis, sensitivity and specificity exhibited reciprocal changes, in the setting of the stable AUC.

Using a parallel machine learning-based approach, we examined which ECG segments contributed most strongly to the prediction of ComVE. The top ECG segments contributing to ECG-prediction of ComVE were related to ventricular depolarization and repolarization, such as the early-mid ST and QRS segments from leads V2-V4, and I, III and aVR. Early-mid portions of PR intervals (from lead I and V5) were also amongst the top 10 segments important for ComVE prediction (**Figure 5, Supplemental Table 1**). Interestingly, after excluding patients with moderate-severe or greater MR from the cohort and re-examineing the ECG segments most strongly contributing to ComVE prediction, both P-wave/PR interval segments (previously in the top 10 strongest ECG-segment predictors) were no longer important predictors (**Supplemental Table 2**).

**Figure 5.**
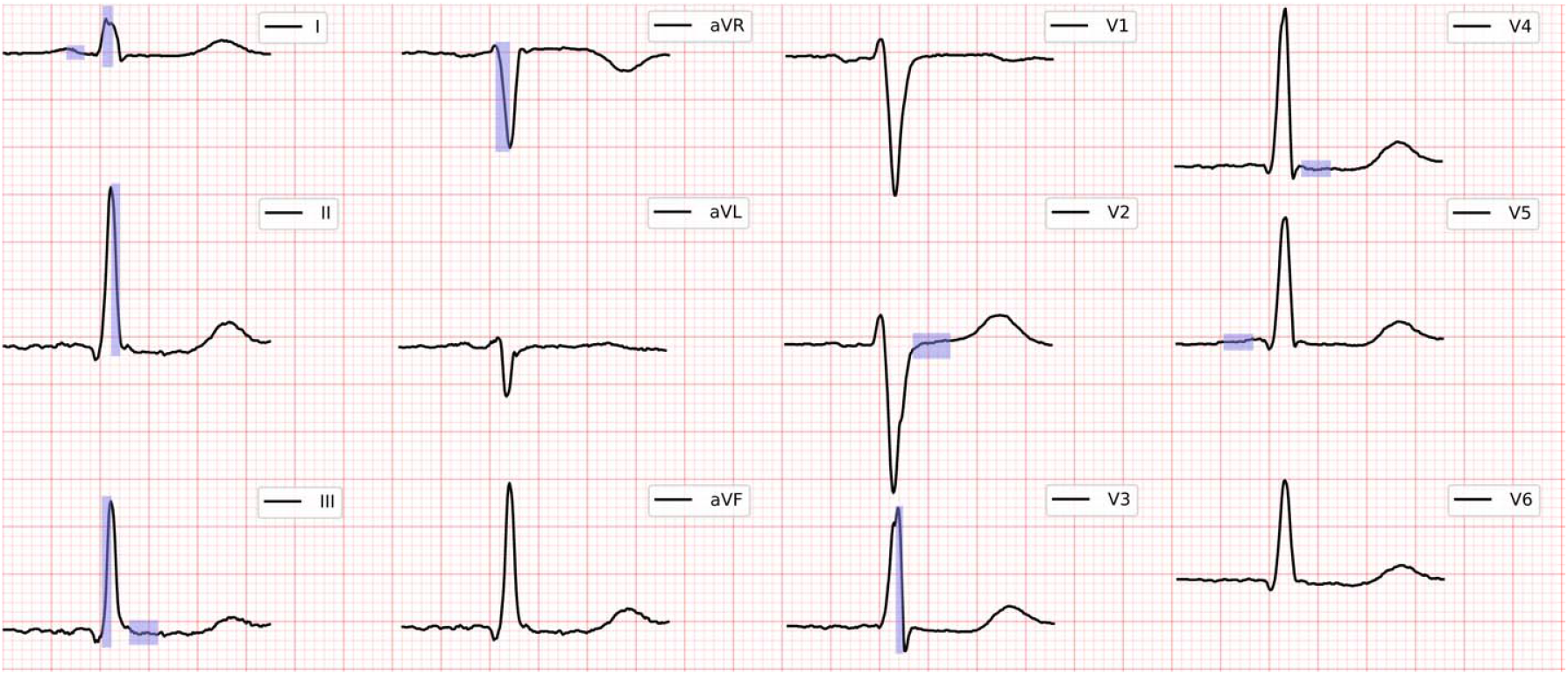
Importance of ECG segments to the prediction of complex ventricular ectopy (ComVE) as determined using a machine learning approach. Highlighted ECG segments indicate the top 10 most important ECG segments for prediction of ComVE in this cohort.

### Performance of a CNN to predict LGE

In a subset of the cohort with CMR data (n=87 patients, 1,369 ECGs), we trained a separate CNN algorithm to discriminate between patients with and without CMR-detected LGE, which represents myocardial scar and predisposes to ventricular arrhythmia (**Figure 6A**).^10, 18^ LGE in the papillary muscles or basal inferolateral wall was present in 21 (24%) of 87 patients with available CMR. Overall, this CNN had an AUC of 0.75 (95% CI, 0.68-0.82) to predict LGE from an ECG, with a sensitivity of 100% and specificity of 54.1% in the hold-out test dataset (**Figure 6B**). For the prediction of LGE, the ECG segments which contributed most strongly were the early-mid QRS from lead I and the early-mid P-wave/PR intervals from V1, V6, II and aVR (**Supplemental Table 3**). When we excluded MVP patients with moderate-severe MR in this cohort, P-wave/PR interval segments were no longer among the most important predictors of LGE. Instead, QRS-related ECG segments showed the greatest importance to predict LGE without moderate-severe MR (**Supplemental Table 4**).

**Figure 6.**
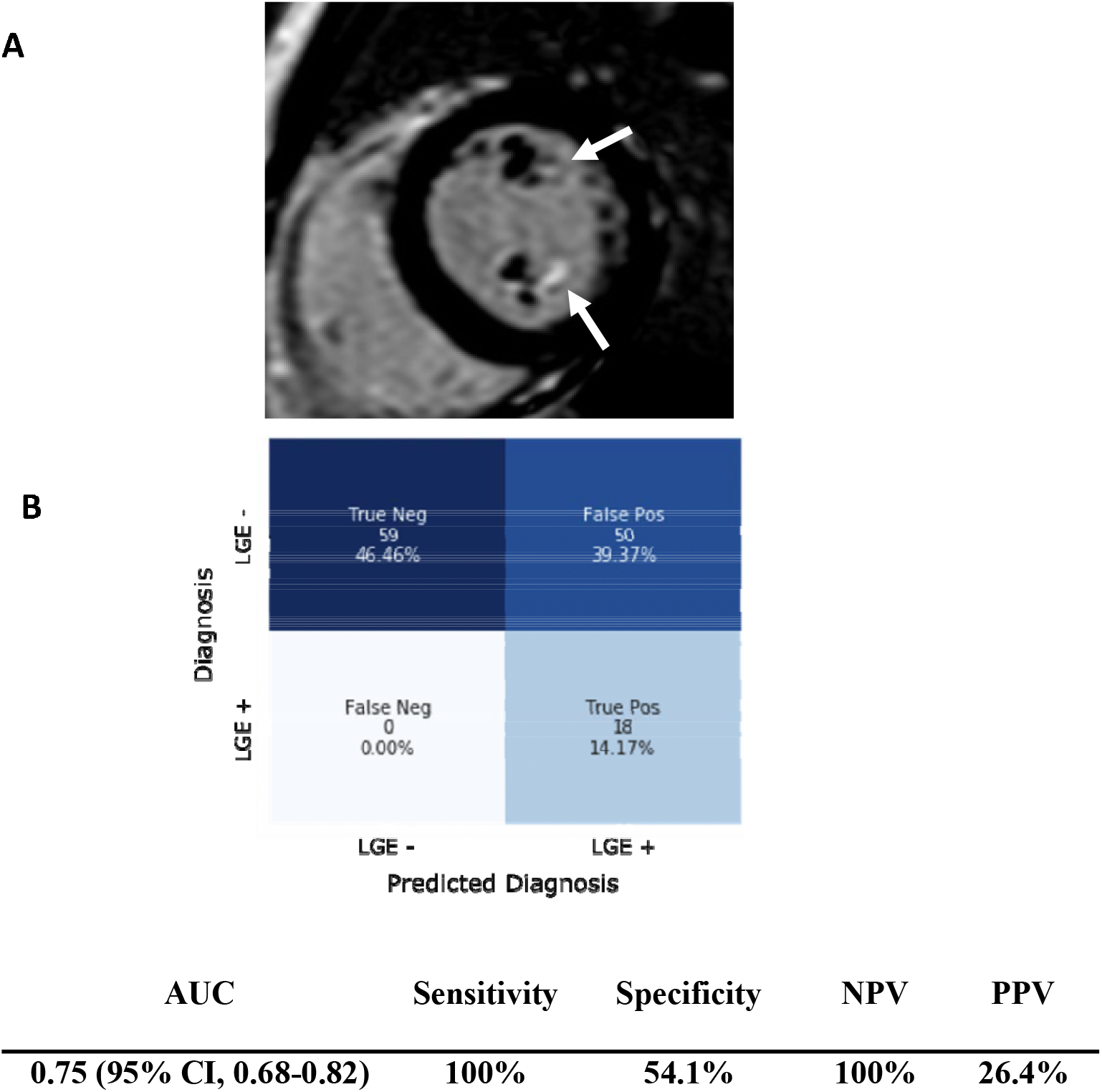
Performance of the ECG-based convolutional neural network (CNN) to detect late gadolinium enhancement (LGE) by cardiac MRI in the test dataset. (A) Magnetic resonance imaging showing late gadolinium enhancement (LGE) in the papillary muscles (arrows). (B) Confusion matrix demonstrating CNN performance to detect LGE with ECG in the LGE test dataset. Performance metrics are shown in the chart (lower). AUC = Area Under the receiver operating characteristic Curve, NPV = Negative Predictive Value, PPV = Positive Predictive Value.

## DISCUSSION

ComVE is detected in most patients with MVP prior to SCA or SCD, is commonly associated with myocardial fibrosis, and is linked to higher all-cause mortality.^5, 10, 24^ In our study, we demonstrate that deep-learning can be applied to standard 12-lead ECGs to 1) identify MVP at risk for ComVE, including those that will develop sustained VT or ventricular fibrillation, 2) identify novel ECG correlates of myocardial disease and arrhythmic risk across mono or bileaflet MVP subtypes, and 3) select those MVPs with CMR-detected myocardial fibrosis. The ability to identify ComVE with an inexpensive, rapid, and widely available point-of-care test such as a 12-lead ECG using deep learning could markedly improve arrhythmic risk stratification in the MVP population. Specifically, MVPs identified to be at risk for ComVE by ECG may benefit from a Holter or event monitor, even if asymptomatic for palpitations. Detection of ComVE at an earlier stage may lead to closer clinical and imaging follow-up, use of more targeted therapies, and the reduction of future severe arrhythmic events.

We have previously shown that deep-learning ECG analysis can discriminate individuals with MVP from those without MVP. However, discrimination for “general” MVP was not as strong as that for arrhythmic MVP (i.e. with ComVE) (AUC 0.77 for MVP^25^ vs AUC 0.81 for ComVE in the current study, **Figure 3**). These findings suggest that ECG features specific to arrhythmic MVP may be absent among mostly benign, non-arrhythmic MVP cases within a large database.

In the ComVE group, 25 individuals (4% of the entire MVP cohort) developed severe ventricular arrhythmia, including high burden of PVCs or VT requiring radiofrequency catheter ablation, sustained VT/ventricular fibrillation treated with an ICD, and even SCD. In such “progressors”, the ComVE CNN score increased over time, in contrast to those MVP with ComVE without progression. These findings suggest that the CNN has increased ability to discriminate over time in those with progressive arrhythmogenicity. Further, this also highlights how the ECG-based neural network may not only function to discriminate ComVE, but may also capture aspects of disease severity or risk of future outcomes such as malignant arrhythmias. Moreover, MVPs with ComVE (either with or without progression) had an early, brief drop in the ComVE CNN score which corresponded to initiation of nodal agents or antiarrhythmics within a similar time-period. Interestingly, the ComVE CNN score increased or stabilized after this drop (**Figure 4C**). Although the observed ComVE CNN score increase is based on a small sample size and warrants further investigation, it may suggest recurrence of arrhythmogenicity and decreased response to medications over time. Indeed, in our sample, MVP patients developed severe arrhythmic events despite use of medications, as previously noted in other studies of MVPs with aborted SCA or SCD, respectively.^5, 10, 20, 32^

There was a greater prevalence of atrial fibrillation or flutter among MVPs with ComVE, suggesting that the higher use of nodal agents or antiarrhythmics in this group may have been partially related to atrial arrhythmias. Concomitant occurrence of atrial and ventricular arrhythmias is common in MVP. Hence, our deep-learning ECG model was developed to reflect true clinical scenarios where use of the same medications may affect the burden of different types of arrhythmia, although prediction of ventricular arrhythmia was the focus of our study.

MVPs without ComVE also had an increase in CNN score over time (**Figure 4C**), albeit consistently below the threshold of 0.39. A mild increase of CNN score may occur in individuals with MVP due to pro-fibrotic conditions such as hypertension or normal aging, although it may never manifest clinically as ComVE.

Ventricular arrhythmias in MVP have been traditionally linked to two echocardiographic phenotypes. In the “hemodynamic” MVP subtype with severe MR, acute or chronic ventricular volume load represents an important arrhythmic trigger,^9^ and even more so if associated with myocardial fibrosis.^17, 20, 33, 34^ More recently, the literature has focused on the “bileaflet” phenotype, which has been linked to increased risk of SCA/SCD even in the absence of significant MR.^5, 10^ In this phenotype, autopsies and CMR with LGE assessment have revealed focal (replacement) fibrosis in the papillary muscles or inferolateral base of the left ventricle (**Figure 5**), as a consequence of abnormal annular mechanics and localized traction on the myocardium.^16, 35^ To investigate the contributions of these two important MVP phenotypes on the CNNs ability to predict ComVE, we did sensitivity analyses excluding patients with a history of MV repair or replacement, moderate-severe or greater MR, or bileaflet MVP. After such exclusions, the AUC remained largely unchanged, suggesting that its overall ability to discriminate ComVE from the ECG occurs independently from the influence of these conditions. These findings highlight that arrhythmogenicity in MVP cannot be attributed only to two echocardiographic phenotypes (significant MR or bileaflet MVP). As observed in true clinical scenarios, there are “gray zones” or combined phenotypes that may be equally important in causing ventricular arrhythmia in MVP. Indeed, only 50% of SCD cases in MVP can be explained by severe MR.^4^ Moreover, in a large matched retrospective cohort study, isolated bileaflet MVP did not appear to significantly increase the risk of SCD/SCA compared with monoleaflet MVP, suggesting the importance of additional risk factors at a population level.^36^ While CNN sensitivity and specificity changed with exclusions of severe MR or bileaflet MVP compared to the primary analysis, changes were reciprocal (i.e. a higher sensitivity and lower specificity or vice versa) and primarily due to the use of the same model threshold as the one used for the primary analysis; overall model performance was unchanged as exhibited by the similar AUC. In clinical or applied settings, the CNN threshold used to identify ComVE could be further tuned in a target population of interest, to optimize (or balance) sensitivity or specificity depending on the task of interest (i.e. screening vs. recruitment for an invasive procedure). Importantly, high negative predictive values could be achieved in both the primary analysis and the sensitivity analyses, which is essential for an ideal diagnostic test.

In our study, we used a novel, purpose-built machine learning approach to examine which ECG segments most strongly contribute to ECG-based ComVE prediction in our cohort, while highlighting potential ECG-associated physiologic underpinnings. The top ECG segments contributing to ComVE prediction were related to ventricular depolarization and repolarization, likely reflecting an underlying MVP-related myopathy that may not be caused only by severe MR (**Supplemental Table 1**). Indeed, when we excluded patients with moderate-severe MR, ECG segments with the highest importance remained the ones related to ventricular activity, whereas the two previously-included P-wave/PR interval-segments were no longer in the top 10 strongest ECG-segment predictors. This is consistent with our understanding of ECG-related cardiac physiology, since longstanding MR has been correlated with atriopathy, which can be reflected by abnormalities of the P wave and PR intervals. In our study, biphasic or inverted T waves in the inferior leads previously described in small, selected samples as a cardinal feature of bileaflet/arrhythmic MVP,^5^ were present in only 17% of patients with ComVE, and in a similar proportion to those without ComVE. Hence, our data-driven machine learning approach provides novel ECG correlates of myocardial disease and arrhythmic risk beyond traditional ECG features of MVP/ComVE, in both mono and bileaflet MVP subtypes. This finding was also corroborated in the LGE subcohort, as discussed below. We believe that machine learning’s potential extends beyond disease prediction and diagnosis tasks, and is ideally positioned to aid in the discovery of biologic and physiologic disease correlates discernable from large-scale medical data to better inform understanding of pathophysiology. By first using machine learning to reveal patterns in large-scale raw ECG data, these findings provide support for future directed investigations into disease mechanisms and physiology. To our knowledge this is one of the first applications of multiple machine learning algorithms to a large cohort of raw ECGs to highlight new physiologic correlates of malignant arrhythmias.

In our cohort LGE was present in the minority of MVPs with ComVE and available contrast CMR, as observed in prior studies.^19, 20^ LGE was more common in those MVPs with ComVE compared to without ComVE, and was localized either in the papillary muscles or basal inferolateral wall. The sensitivity and negative predictive value of our ECG-based deep-learning model for predicting LGE were excellent (100%), highlighting the potential usefulness of this screening tool in a clinical setting. We have shown both in living individuals and in post-mortem samples, that MVPs with ventricular arrhythmia or SCD do not consistently have replacement fibrosis.^20, 23^ In contrast, diffuse interstitial fibrosis is common, with and without significant MR. Diffuse fibrosis (either primary or MR-related), may cause ECG abnormalities even in the absence of LGE, thus reducing the specificity of our CNN. The ECG segments which contributed most strongly to the LGE prediction were related to ventricular activity, possibly reflecting an underlying MVP-related myopathy. However, atrial activity also affected LGE prediction, likely reflecting the contribution of severe MR to development of LGE (**Supplemental Table 2**).^17^ Interestingly, when we excluded patients with moderate-severe or greater MR, P-wave and PR-interval ECG segments ceased to be among the top 10 strongest ECG-segment predictors of LGE, which instead were all QRS-related. Similar to our finding for ComVE, there may be ECG changes associated with a primary, MVP-related myopathy^6, 20, 37^ that enable ECG-based prediction of LGE even in the absence of significant MR. Deep learning analysis of ECGs may provide a more cost-effective way to select candidates for a CMR to detect fibrosis when traditional arrhythmic echocardiographic phenotypes such as bileaflet MVP or severe MR are absent.

### Limitations

First, our study was not sufficiently powered to develop an ECG-based CNN to predict SCD or SCA. However, we believe studying an intermediate stage of disease (ComVE) before development of SCA may be equally important for the purpose of risk stratification, an unmet need in the MVP population, which comprises mostly benign cases. Similarly, our sample size was not sufficiently large to determine whether those identified by the CNN as having ComVE would develop SCA or SCD in the future. However, we did identify higher ComVE CNN scores over time in those MVPs that progressed to more severe ventricular arrhythmia (including SCA/SCD) compared to those that did not progress. These findings suggest that CNNs could potentially identify those at risk for SCD/SCA from ECGs alone if applied to a larger study sample. We theorize that in some patients our LGE CNN algorithm may have detected ECG features related to interstitial/diffuse rather than focal fibrosis. ^19, 20 23 37^ However, we could not confirm this observation, as T1 mapping was not available in all patients with a CMR.

## CONCLUSIONS

A CNN can detect MVP patients at risk for both ventricular arrhythmias and CMR-measured fibrosis from standard 12-lead ECGs, and can identify novel ECG correlates of arrhythmic risk regardless of leaflet involvement or MR severity. Deep learning-based analysis of ECGs may help identify, within a large database, those MVP patients requiring closer follow-up and/or a CMR.

## Data Availability

The data used in this study are derived from clinical care and thus are not made publicly available due to data privacy concerns. Reasonable requests for collaboration using the data can be made from the authors, as feasible and permitted by the Regents of the University of California.

## SOURCES OF FUNDING

This work was supported by the UCSF Cardiology Innovation Award and by the National Institutes of Health NHLBI R01HL153447 (Dr Delling) and NHLBI K23HL135274 (Dr. Tison). The funders had no role in design and conduct of the study; collection, management, analysis, and interpretation of the data; preparation, review, or approval of the manuscript; and decision to submit the manuscript for publication.

### Code Availability

The code that supports this work is copyright of the Regents of the University of California and can be made available through license.

